# The Prevalence of Self-reported ADHD among University Students in Jordan

**DOI:** 10.64898/2026.05.29.26354419

**Authors:** Omar al-Omoush, Sara M. Farah, Mishkat Ihsan, Rama Al-Safadi, Yanal Aldaoud, Layan Alali, Leen M. Ahmed, Ayoub Al-Hijazin, Suleiman Shahatit, Hamzeh Al-Shenag, Abdullah Alseidi

**Affiliations:** Department of Family Medicine, Faculty of Medicine, Hashemite University, Zarqa, Jordan; Faculty of Medicine, Hashemite University, Zarqa, Jordan

## Abstract

**Background:** Attention Deficit Hyperactivity Disorder (ADHD) is characterized by persistent inattention, hyperactivity, and impulsivity. While documented in children, research on its persistence into young adulthood in Jordan remains scarce. This gap is critical given the cognitive demands of higher education. This study estimated attention deficit hyperactivity disorder (ADHD) symptom prevalence among Jordanian university students, examined associations with gender and academic performance, and identified barriers to mental health service accessibility.

**Methods:** A descriptive cross-sectional study using web-based sampling recruited 389 university students (aged ≥ 18 years) from various Jordanian universities. Participants completed an online survey, incorporating the validated English and Arabic versions of the Adult ADHD Self-Report Scale (ASRS-v1.1) to assess symptom prevalence, alongside inquiries regarding demographics, academic history, and barriers to care.

**Results:** The prevalence of probable ADHD was 37.5% (n=146). Males constituted a significantly higher proportion of positive cases (69.9%) compared to females (30.1%). A strong statistical association was found between positive ADHD screening and negative academic impact (p<0.001), as well as negative effects on emotional well-being (p<0.001). Comorbidities including anxiety disorders and emotional abuse were significantly linked to probable ADHD (p=0.019). Notably, positive-screened participants were significantly more likely to cite social stigma as a primary barrier to seeking professional help (p=0.024).

**Conclusion:** Self-reported ADHD symptoms are highly prevalent among Jordanian university students, correlating with substantial academic underachievement and emotional dysfunction. These findings highlight an urgent need for targeted university-based screening programs, academic accommodations, and de-stigmatization campaigns to facilitate early intervention and improve educational outcomes in this population.

## 1. Introduction and Background

Attention-deficit hyperactivity disorder (ADHD) is a chronic, neurologically based illness characterized by a persistent pattern of inattention and/or hyperactivity and impulsivity that are more inappropriate or disruptive than those in other children of a comparable age [1]. Globally, studies estimate that ADHD affects between 5% and 7.2% of youth and between 2.5% and 6.7% of adults [2,3,4]. ADHD is not merely a disorder that children “eventually grow out of” as some members of society believe, leading to a high prevalence of undiagnosed cases [5]. Research has shown that the screening, diagnosis, and documentation of ADHD can vary significantly and are influenced by various factors, including the impact of stigma [6]. Some studies point out that stigma is deeply embedded in the familial and socio-cultural fabric of Arab society, leading to the persistent and blatant violation of the fundamental rights of individuals with mental illness. This issue is likely to intensify as the prevalence of mental disorders increases in the region, driven by ongoing political and economic instability [7]. This issue can adversely impact the course of mental illness among Arabs by delaying accurate diagnosis and appropriate treatment [8]. Awareness campaigns have a significant role in combating mental health stigma by normalizing seeking help and motivating individuals to take action when they need support [9,10]. Hence, shedding light on the importance of accurate diagnosis and raising awareness about available support resources, can help address the stigma surrounding ADHD and promote a more informed and supportive environment for those affected especially in universities.

Acknowledging that ADHD can significantly impair an adult’s quality of life, especially if the disorder goes unrecognized, is crucial. Early and accurate diagnosis, coupled with appropriate interventions, can substantially enhance long-term outcomes for individuals with ADHD [11].

### 1.1. Adult ADHD

Attention-deficit/hyperactivity disorder (ADHD) in adulthood remains a complex and evolving area of research. Symptom presentation changes with age, as hyperactivity and impulsivity tend to decline more rapidly than inattention, which often persists into later life [11]. Longitudinal studies indicate that ADHD frequently continues into adulthood, with approximately 60% of affected individuals exhibiting either full diagnostic criteria or residual symptoms, often resulting in a chronic clinical course [12]. While most evidence supports the persistence of ADHD from childhood, emerging data suggest that ADHD may also manifest in adulthood in individuals without clear childhood symptoms [13].

In such cases, individuals may have experienced mild or subclinical symptoms during childhood within relatively low-demand environments. However, exposure to increased cognitive, social, and occupational demands in adulthood, such as university education, competitive employment, or family responsibilities may exacerbate these symptoms, leading to functional impairment and eventual clinical recognition [14]. Therefore, increasing awareness of adult ADHD among individuals transitioning into higher education and professional settings is essential.

Using the Adult ADHD Self-Report Scale (ASRS), a study conducted in Saudi Arabia reported that 11.9% of college students met diagnostic criteria for ADHD, while another study in the United Arab Emirates found that 34.7% of students reported ADHD symptoms [6, 15].

Adult ADHD is frequently associated with psychiatric and medical comorbidities, which may complicate diagnosis and adversely affect disease progression [12]. This highlights the importance of early identification and the establishment of structured support systems for affected university students.

Despite growing evidence that ADHD persists beyond childhood, adult ADHD continues to be under-recognized. Many affected individuals remain undiagnosed due to limited clinician awareness and insufficient adherence to established diagnostic guidelines [12]. Furthermore, social stigma, particularly from family members, may discourage individuals from seeking evaluation and treatment [14].

The personal, social, and economic burden of untreated adult ADHD is substantial and can significantly compromise quality of life [11]. Therefore, improved recognition, timely diagnosis, and effective management of adult ADHD are essential to enhance individual functioning, productivity, and overall well-being.

### 1.2. Influences on ADHD

Attention-deficit/hyperactivity disorder (ADHD) is influenced by multiple interacting factors, including genetic and familial predisposition, environmental exposures, socioeconomic conditions, parental practices, and gender.

#### 1.2.1 Genetic and familial factors

ADHD is recognized as both familial and highly heritable, with genetic and molecular studies demonstrating a substantial overlap between ADHD and other neurodevelopmental conditions, particularly autism spectrum disorders [16,17]. Consanguinity has also been linked to an increased risk of ADHD, with one study reporting that 76% of patients with ADHD had a history of parental consanguinity [18].

#### 1.2.2 Family, Socioeconomic Status, and Parental Practices

Socioeconomic factors and parenting dynamics play a critical role in the prevalence and management of ADHD. Children from lower socioeconomic backgrounds are 1.85–2.21 times more likely to receive an ADHD diagnosis, an association often attributed to increased exposure to environmental risks, such as nutritional deficiencies and limited healthcare access [19].

Additionally, maternal employment status and specific parenting practices—including frequent punishment, poor monitoring, and unsupportive strategies—have been linked to the persistence of symptoms over time [20–22].

#### 1.2.3 Gender differences

Historically, ADHD was viewed as a predominantly male disorder; however, contemporary evidence suggests that females are affected at comparable rates, as they more frequently present with inattentive symptoms rather than hyperactivity [23].

### 1.3. ADHD Diagnosis

Screening for ADHD in adults is essential, as untreated ADHD can significantly affect personal, professional, and social functioning [24]. Many adults with ADHD remain undiagnosed from childhood, resulting in prolonged difficulties that interfere with daily life and overall productivity [14]. Comprehensive screening and structured assessment in adulthood facilitate accurate identification of ADHD and improve access to appropriate diagnosis, treatment, and long-term support.

The diagnostic framework for ADHD has evolved substantially, with the Diagnostic and Statistical Manual of Mental Disorders (DSM) serving as the primary reference standard. The Diagnostic and Statistical Manual of Mental Disorders - Fourth Edition (DSM-IV), published in 1994, defined diagnostic criteria based on symptoms of inattention, hyperactivity, and impulsivity [25]. However, these criteria were largely derived from pediatric populations, which limited their applicability to adult presentations of ADHD.

The introduction of the Diagnostic and Statistical Manual of Mental Disorders – Fifth Edition (DSM-V) in 2013 addressed this limitation by expanding diagnostic criteria to better reflect adult symptom patterns. The DSM-V acknowledges that ADHD may persist into adulthood and may present differently, with adults frequently experiencing impairment in social, academic, and occupational domains [26]. Early and accurate diagnosis enables improved symptom recognition, timely treatment, and the development of effective coping strategies, emotional regulation skills, and functional adaptation, ultimately enhancing quality of life.

Despite the well-established adverse outcomes associated with untreated ADHD, relatively few studies have focused on the consequences of undiagnosed ADHD and its associated comorbidities [27].

### 1.4. Effects of ADHD

ADHD is associated with a wide range of adverse effects that extend beyond the affected individual to impact family functioning [28], peer relationships from childhood into adulthood [29], and later life outcomes, including financial stability and job attainment [30]. These cumulative challenges highlight ADHD as a disorder with substantial lifelong personal and societal consequences.

#### 1.4.1. Psychiatric disorders and comorbidities

A diagnosis of ADHD should prompt careful evaluation for psychiatric comorbidities, as comorbid disorders are highly prevalent in this population. Owing to the frequent underdiagnosis of ADHD, routine screening is recommended for patients presenting with multiple psychiatric conditions, including substance use, mood, and anxiety disorders. Meta-analytic evidence indicates that substance use disorder (SUD) is the most common comorbidity, with approximately one in four adolescents and adults with substance abuse disorder (SUD) meeting criteria for ADHD [31–32]. This is followed by mood disorders, anxiety disorders, and personality disorders, particularly antisocial personality disorder (ASPD) [33].

Research has established shared neurobiological mechanisms between ADHD and substance abuse disorder (SUD), including dopaminergic dysregulation of reward and motivational pathways [33]. A study conducted in Lebanon further demonstrated that individuals with ADHD experienced an earlier onset of SUD, longer durations of substance abuse, and relapse rates three times higher than those observed in individuals with SUD without ADHD [34]. These findings emphasize the clinical importance of early ADHD identification in populations with substance-related disorders.

Bipolar disorder also frequently co-occurs with ADHD, with approximately 33% of youth with ADHD reported to have comorbid bipolar disorder [35–36].

#### 1.4.2. Effect on sleep

Sleep disturbances represent a major concern in individuals with ADHD, with a high prevalence of insomnia-related symptoms reported in the literature. Approximately 80% of untreated patients with ADHD have been reported to experience symptoms of sleep-onset insomnia [37]. It is important, however, to distinguish between the presence of insomnia symptoms and a formal diagnosis of insomnia disorder. One study found that 44.4% of patients met DSM-5 criteria for insomnia disorder, while 63.9% reported insomnia symptoms without necessarily meeting diagnostic criteria [38]. Overall, the prevalence of insomnia symptoms in ADHD populations has been estimated to range from 43% to 80% [39–41].

ADHD severity has been shown to be independently associated with insomnia disorder [38], highlighting the negative impact of both hyperactivity/impulsivity and inattentive symptoms on sleep initiation and maintenance [42–43]. These findings suggest a complex and potentially bidirectional relationship between ADHD symptom severity and sleep impairment.

The importance of treatment is further underscored by evidence indicating that patients receiving pharmacological therapy tend to demonstrate a lower prevalence of insomnia disorder. Moreover, longer durations of stable treatment have been associated with reduced rates of both insomnia disorder and insomnia symptoms [38].

#### 1.4.3. Effect on daily life, social interactions, family bonds and relationships

Adults with ADHD frequently experience unstable or strained relationships with their partners and report a reduced perception of their ability to provide emotional support. Poor listening skills and frequent interruptions further impair effective communication and conflict resolution, negatively affecting relationship quality [11].

In addition to emotional challenges, adults with ADHD experience significant functional impairments in daily life. They report greater difficulty managing heavy academic workloads, maintaining concentration during studies, and organizing assignments. Moreover, they are less likely to be employed in full-time positions, and those who are employed tend to change jobs more frequently over a ten-year period [11].

#### 1.4.4. Smoking and caffeine consumption

Previous research demonstrates a significant association between ADHD and increased rates of smoking and caffeine consumption. This relationship has been attributed to alterations in dopaminergic regulation in individuals with ADHD, as dopamine plays a central role in attention, motivation, and reward processing. Such dysregulation may predispose individuals with ADHD to behaviors that stimulate the dopamine reward pathway, including nicotine use [44].

Nicotine has been shown to influence executive dysfunction, inattention, attentiveness, and both cognitive and behavioral inhibition, core features of ADHD. Consequently, many individuals with ADHD may engage in nicotine use as a form of self-medication, despite the well-established risks of nicotine dependence [45–46]. A 2017 study reported that nicotine improved attentiveness, reduced inattention, enhanced executive functioning, and improved cognitive and behavioral inhibition in individuals with ADHD, providing further explanation for the higher prevalence of nicotine use in this population compared with the general population [46].

Similarly, caffeine is another commonly consumed stimulant among individuals with ADHD due to its modest dopaminergic effects. However, excessive caffeine intake may contribute to dependence and exacerbate anxiety and sleep disturbances, although these effects are generally less pronounced than those associated with nicotine use [47].

#### 1.4.5. Academic performance

Academic performance has been extensively studied in relation to various psychological disorders, including Attention-Deficit/Hyperactivity Disorder (ADHD) [48–49]. It represents a critical determinant of children’s educational achievement and future opportunities. Numerous studies have demonstrated a strong association between ADHD and academic underachievement, as well as poorer educational outcomes [50–52].

The three core symptoms of ADHD—impulsivity, hyperactivity, and inattention—can substantially impair an individual’s ability to sustain attention, regulate behavior, and complete academic tasks efficiently. These difficulties often result in lower academic grades, increased dropout rates, and problems with task completion and time management [50].

Although treatments for ADHD have been shown to improve academic performance, their effectiveness varies depending on treatment modality and individual patient characteristics [53].

#### 1.4.6. Domestic Violence

Studies have consistently demonstrated that children with Attention-Deficit/Hyperactivity Disorder (ADHD) are at a higher risk of exposure to physical, psychological, and emotional abuse, as well as neglect, compared to their non-ADHD peers [54]. This increased vulnerability is thought to arise from a combination of factors, including the behavioral challenges associated with ADHD symptoms, the heritable nature of the disorder, and broader family and social dysfunction. In childhood, symptoms such as emotional dysregulation, temper tantrums, restlessness, and physical or verbal aggression may place additional strain on caregivers and family systems [54].

Previous research has also shown that young women with a history of childhood ADHD are at an increased risk of experiencing physical intimate partner violence (IPV) in adulthood. Greater ADHD symptom severity has been linked to higher intimate partner violence (IPV) risk, potentially mediated by factors such as academic underachievement, risky sexual behaviors, and the presence of internalizing (e.g., depression, anxiety, low self-esteem) and externalizing (e.g., conduct disorder, oppositional defiant disorder) symptoms during adolescence and young adulthood [55].

The hereditary nature of ADHD suggests that affected children may have parents who also experience adult ADHD or other mental health disorders. These parental challenges may contribute to heightened stress, inconsistent parenting practices, and family dysfunction, further increasing the risk of adverse childhood experiences [54].

These findings highlight the complex relationship between ADHD and adverse interpersonal experiences and underscore the importance of targeted prevention and intervention strategies, particularly for vulnerable populations. Academic support and empowerment may play a crucial protective role in mitigating long-term adverse outcomes, including intimate partner violence, among individuals with ADHD [55].

### 1.5. Treatment

Despite the availability of effective treatment options, ADHD remains substantially undertreated in adulthood, with only approximately 11% of adults with an ADHD diagnosis receiving treatment [56]. Current management strategies are broadly categorized into pharmacological and non-pharmacological approaches, with medication-based therapy remaining the most utilized intervention [57].

#### 1.5.1. Pharmacological Treatment

Pharmacological treatment primarily consists of psychostimulants, including methylphenidate-based agents (e.g., Ritalin) and amphetamine-based medications (e.g., Adderall), as well as non-stimulant alternatives [27]. Psychostimulants are generally considered first-line therapy due to their well-established efficacy, achieving symptom improvement in up to 70% of patients [56]. However, concerns regarding adverse effects and misuse contribute to ongoing debate surrounding their use [27]. For individuals with inadequate response to pharmacotherapy alone, combined pharmacological and non-pharmacological treatment has demonstrated superior outcomes [57].

#### 1.5.2. Non-Pharmacological Treatment

Non-pharmacological interventions, particularly psychotherapy and cognitive-behavioural therapy (CBT), have shown meaningful benefits in adults with ADHD [57]. Although these approaches may not directly target core neurobiological symptoms, they effectively address functional impairments related to academic, occupational, and interpersonal difficulties by enhancing organizational skills, time management, problem-solving, and self-regulation [57]. Mindfulness-based interventions, including meditation and yoga, are increasingly employed as adjunctive therapies and have been associated with improved cerebral blood flow and glucose metabolism in brain regions involved in attention and emotional regulation, such as the prefrontal and cingulate cortices [57].

### 1.6. Stigma

Mental health stigma refers to social disapproval, disgrace, or discrediting of individuals with mental health conditions and remains a major barrier to diagnosis, treatment, and recovery [58–59]. Stigma arising from broader societal attitudes toward individuals perceived as behaviorally or cognitively different can delay help-seeking, worsen existing symptoms, and restrict access to essential life opportunities, including education, employment, healthcare, and housing [60].

Mental and behavioral disorders are particularly stigmatized in many cultures, including the Arab region and the Middle East. A systematic review of stigma related to mental illness in Arab societies identified wide variability in stigmatizing beliefs, attitudes, and behaviors toward individuals with mental health conditions and their treatment [6,61]. In some Arab countries, stigma is further reinforced at the policy level, where limited resource allocation and inaccurate portrayals of mental illness contribute to systemic barriers to care [8]. In Jordan, analysis of mental health clinic databases revealed that stigma accounted for 41% of delayed visits, treatment discontinuation, noncompliance, and delayed clinical improvement, highlighting the substantial impact of public stigma on mental health outcomes, particularly among younger individuals [62]. Additionally, studies on mental health literacy in Jordan demonstrate that cultural norms and familial expectations significantly shape mental health perceptions and behaviors, further emphasizing the influence of sociocultural factors on care-seeking [63].

Collectively, these findings underscore the persistence of stigmatization within both the community and healthcare settings [7].

Thus emphasizing the importance of proactive engagement by healthcare professionals in addressing mental health stigma through patient education and timely psychological and pharmacological interventions [62]. Broader strategies, including integrating mental health education into academic curricula, implementing public media campaigns to challenge stigma, and embedding mental health services within general healthcare systems, are essential to improving mental health awareness and access to care in Arab populations [8].

## 2. Methods

### 2.1. Study Design and Participants

A descriptive cross-sectional study using web-based convenience sampling was conducted among 389 university students aged ≥18 years from various universities in Jordan. Participants were recruited through an online survey distributed across student networks and social media platforms. The questionnaire was completed anonymously and included items assessing demographic characteristics, academic history, and perceived barriers to seeking care.

### 2.2. Instrument

The prevalence of adult attention-deficit/hyperactivity disorder (ADHD) symptoms was assessed using the validated English and Arabic versions of the Adult ADHD Self-Report Scale – Version 1.1 (ASRS-v1.1). The ASRS-v1.1 is a widely used screening tool based on diagnostic criteria outlined in the Diagnostic and Statistical Manual of Mental Disorders Fourth Edition (DSM-IV). The full scale consists of 18 items evaluating ADHD symptoms experienced during the previous six months, with responses recorded on a five-point Likert scale ranging from “never” to “very often.”

Six of the 18 items were identified as the most predictive of ADHD and form the short-version screener used in this study. This shortened version demonstrates strong psychometric properties, with a reported sensitivity of 68.7% and specificity of 99.5%, an overall classification accuracy of 97.9%, and high internal consistency [64–65].

The survey also collected demographic information including age, gender, academic year, field of study, academic performance, and relevant medical history. Additional items assessed parental educational level and family history of mental health conditions to explore potential familial associations.

### 2.3. Study Procedure and Statistical Analysis

Collected data were entered and analyzed using IBM SPSS Statistics. Responses with incomplete data were excluded from the analysis. Descriptive statistics were used to summarize sociodemographic characteristics. Associations between categorical variables were assessed using the chi-square test or Fisher’s exact test when appropriate. A p-value of <0.05 was considered statistically significant, and 95% confidence intervals (CIs) were reported where applicable.

### 2.4. Ethical Approval

Ethical approval for this study was obtained from the Institutional Review Board (IRB) of The Hashemite University on January 1, 2025. Data collection commenced following Institutional Review Board (IRB) approval, and continued until June 1, 2025, at which point survey responses were accessed for analysis. Participation in the study was voluntary, and informed consent was obtained electronically from all participants prior to completing the questionnaire. All responses were collected anonymously and were used solely for research purposes.

## 3. Results

### 3.1. Participant Characteristics

A total of 389 university students in Jordan participated in the study. Among them, 62.5% (n = 243) screened negative for ADHD, while 37.5% (n = 146) met the ASRS-v1.1 criteria for probable ADHD. Only 6.7% (n = 26) reported a prior formal diagnosis of ADHD. The mean age of participants was 21 ± 1.5 years, with a slightly higher proportion of males among those screening positive (69.9%, n = 102) compared to females (30.1%, n = 44).

A similar distribution was observed among participants reporting a prior formal diagnosis of ADHD (69.2% male vs. 30.8% female). However, despite this higher proportion of males in the diagnosed group, the gender difference for previously established diagnoses did not reach statistical significance within this sample.

### 3.2. Family History and Socioeconomic and parenting practices

Among participants with positive ASRS, family histories included anxiety disorders (16.4%), sleep disorders (10.3%), and specific learning disorders (3.4%) (Table 1). And participants with a prior ADHD diagnosis, family histories were more pronounced for mood disorders (57.7%), anxiety disorders (26.9%), and autism spectrum disorders (11.5%) (Table 2).

**Table 1:** Family History in Participants Screening Positive on ASRS.

| Family History Domain | % Positive |
| --- | --- |
| Anxiety Disorders | 16.4% |
| Sleep Disorders | 10.3% |
| Specific Learning Disorders | 3.4% |

**Table 2:** Family History in Participants with ADHD Diagnosis.

| Family History Domain | % Positive |
| --- | --- |
| Mood Disorders | 57.7% |
| Anxiety Disorders | 26.9% |
| **Autism Spectrum Disorders** | 11.5% |

Parental relationship quality was assessed on a five-point scale, ranging from **very strong and supportive (1)** to **very weak or disconnected (5)** showed significant association with ASRS screening outcomes (χ²(4, N = 389) = 27.15, p < 0.001). Participants with positive ASRS results reported lower rates of “very strong” parental relationships (33.6%) compared with “fairly strong” (34.2%), “neutral” (19.2%), “somewhat strained” (10.3%), and “very weak/disconnected” (2.1%) dynamics (Table 3). No significant association was observed between parental relationship quality and formal ADHD diagnosis. Among other investigated genetic and familial factor, consanguinity was statistically insignificant in our study

**Table 3:** Parental Relationship Quality by ASRS Screening Results.

| **Relationship Quality** | **% Positive ASRS** |
| --- | --- |
| **Very Strong** | 33.6% |
| **Fairly Strong** | 34.2% |
| **Neutral** | 19.2% |
| **Somewhat Strained** | 10.3% |
| **Very Weak / Disconnected** | 2.1% |

### 3.3. Psychiatric Comorbidities and Sleep disorders

Participants with anxiety disorders were more likely to screen positive on the ASRS-v1.1 than those without such a history. A statistically significant association was observed between positive ADHD screening and a history of anxiety disorders (p = 0.019). These findings align with previous literature reporting high comorbidity between anxiety disorders and ADHD, which may be attributed to overlapping symptoms such as inattention, restlessness, and executive dysfunction.

A statistically significant association was also observed between a prior diagnosis of ADHD and comorbid mood disorders, with 57.7% of participants with ADHD reporting a history of mood disorders. In contrast, participants who screened positive on the ASRS did not demonstrate a statistically significant association with mood disorders, although screening positivity was slightly higher among individuals with mood disorders (41.2%) compared to those without (37.4%).

A weak but potential association was observed between sleep disorders and prior ADHD diagnosis (p = 0.065). Participants with sleep disorders were more likely to report a previous ADHD diagnosis (14.7%) compared with those without sleep disorders (5.9%). A major limitation of the present analysis is the small number of participants with a history of sleep disorders (n = 34), which limits statistical power and may reduce the ability to detect significant associations. Future studies should include larger samples and incorporate objective sleep assessments to better clarify the direction and mechanisms underlying this relationship

### 3.4. ADHD Effects on Daily Life and Emotional Well-Being

Emotional well-being was significantly associated with ADHD symptomatology as assessed by the ASRS-v1.1. Participants who screened positive for ADHD symptoms were substantially more likely to report negative effects on their emotional health (53.4%) compared with those who screened negative (19.3%), suggesting that a greater ADHD symptom burden may be linked to increased emotional difficulties. Similarly, participants with a self-reported history of ADHD diagnosis reported markedly poorer emotional well-being, with 76.9% (n = 20) indicating a negative impact on their emotional health, while only 23.1% (n = 6) reported no negative impact. Together, these findings indicate a consistent relationship between ADHD diagnosis, higher symptom severity, and poorer emotional well-being, highlighting the potential importance of targeted interventions aimed at improving emotional regulation among individuals with ADHD.

### 3.5. Academic Performance

A statistically significant association was identified between ADHD and academic performance using two complementary measures. First, a self-reported history of ADHD diagnosis was significantly associated with negative academic impact (p < 0.001). This association was even stronger when current ADHD symptoms were assessed using the ASRS-v1.1, yielding a markedly higher Pearson Chi-Square value (χ² = 61.908, p < 0.001), supported by both Likelihood Ratio and Fisher’s Exact Test results.

### 3.6. Smoking, Vaping, and Caffeine Use

Among ASRS-positive participants, 20.6% reported vaping, whereas 42.3% of participants with a prior ADHD diagnosis reported vaping. Chi-square analysis demonstrated a significant association between ADHD diagnosis and vaping (χ² = 8.063, p = 0.005) and smoking (χ² = 11.886, p = 0.001),), which was supported by Likelihood Ratio and Fisher’s Exact Test results, but no significant associations were observed between ASRS screening status and nicotine use. Caffeine consumption was not significantly associated with either ADHD diagnosis or ASRS status (ρ = 0.083, p = 0.067). These associations between nicotine and caffeine consumption and ADHD status are summarized in (Table 4).

**Table 4:** Association between nicotine use and ADHD diagnosis / symptom screening (Pearson Chi-Square Test)

| | Result ($\chi^2$) | p-value | Significance |
| --- | --- | --- | --- |
| Vaping vs. ADHD Dx | 8.063 | 0.005 | Significant |
| Smoking vs. ADHD Dx | 11.886 | 0.001 | Significant |
| Vaping vs. ASRS (+/-) | – | 0.995 | Non-significant |
| Smoking vs. ASRS (+/-) | 0.282 | 0.595 | Non-significant |

### 3.7. Domestic Violence

Emotional abuse was significantly associated with ADHD diagnosis (38.5%, χ² = 5.462, p = 0.019) and ASRS-positive screening (30.8%, p < 0.001). No significant associations were found for physical or sexual abuse.

Among participants who preferred not to answer abuse-related questions, 7.7% of those with an ADHD diagnosis and 11% of those who screened positive on the ASRS did not provide responses for any of the three abuse types

### 3.8. Treatment and Support

Participants with prior ADHD diagnosis were significantly more likely to report medication use (p < 0.001), psychotherapy (χ² = 4.868, p = 0.027), academic support (χ² = 10.019, p = 0.002), and support from family/friends (χ² = 8.609, p = 0.003). Similarly, ASRS-positive participants showed significant associations with counseling/psychotherapy (χ² = 12.528, p < 0.001), medication use (χ² < 0.001), academic support (χ² = 28.840, p < 0.001), lifestyle modifications (χ² = 20.811, p < 0.001), support from family/friends (χ² = 14.830, p < 0.001), and self-help resources (χ² = 4.260, p = 0.039) (Tables 5 and 6).

**Table 5:** Treatment and Support Associated with ASRS-Positive Participants.

| **Intervention /** | **Significant?** | **p-value** |
| --- | --- | --- |
| **Medication Use** | Yes | <0.001 |
| **Counseling / Psychotherapy** | Yes | <0.001 |
| **Academic Support** | Yes | <0.001 |
| **Support from Family & Friends** | Yes | <0.001 |
| **Compensation / Rehabilitation** | No | — |
| **Lifestyle Modifications** | Yes | <0.001 |
| **Self-help Resources** | Yes | 0.039 |

**Table 6:** Treatment and Support Associated with ADHD Diagnosis.

| **Intervention** | **Significant?** | **p-value** |
| --- | --- | --- |
| **Medication Use** | Yes | <0.001 |
| **Psychotherapy** | Yes | 0.027 |
| **Academic Support** | Yes | 0.002 |
| **Support from Family & Friends** | Yes | 0.003 |
| **Compensation / Rehabilitation** | Yes | <0.001 |
| **Lifestyle Modifications** | No | — |
| **Self-help Resources** | No | — |

### 3.9. Perceived Stigma

In the current study, a significant relationship was observed between current ADHD symptomatology and perceived stigma. Participants who screened positive on the ASRS were more likely to report social stigma as a barrier to help-seeking (11.6%, 17/146). This association was confirmed using Pearson’s Chi-square test (p=0.024) and supported by Fisher’s Exact Test (p=0.031), indicating that individuals experiencing active ADHD symptoms perceive greater social obstacles to accessing care.

However, no significant association was found between a formal history of ADHD diagnosis and reporting social stigma as a barrier to seeking help, suggesting that prior clinical diagnosis alone does not significantly influence stigma perception.

## 4. Discussion

This study highlights the multifactorial nature of ADHD among university students, emphasizing the interplay between several factors. Consistent with existing literature, our findings reinforce the strong familial aggregation of ADHD and its overlap with other psychiatric and neurodevelopmental conditions, particularly anxiety disorders, sleep disturbances, autism spectrum disorders, and mood disorders.

Gender differences were evident at the symptom level, with males more likely to screen positive on the ASRS-v1.1; however, this disparity was not statistically significant for self-reported ADHD diagnosis. This finding aligns with growing evidence that ADHD in females may be underdiagnosed due to a more subtle symptom presentation and compensatory strategies, particularly in academic settings.

ADHD was associated with substantial functional impairment across multiple domains. Academic performance showed one of the strongest associations, particularly when ADHD was assessed using current symptom screening rather than prior diagnosis. This highlights the importance of active symptom burden, rather than diagnostic history alone, in predicting real-world impairment. Emotional well-being was also markedly affected, with individuals with ADHD nearly three times more likely to report negative emotional outcomes, underscoring the need for integrated mental health support.

Psychiatric comorbidity patterns in this study were broadly consistent with prior research. Anxiety disorders demonstrated a significant association with ADHD symptoms, whereas mood disorders did not reach statistical significance, suggesting that symptom overlap, self-report bias, and contextual factors may influence observed relationships.

Lifestyle behaviors revealed a nuanced pattern. While caffeine intake was not significantly associated with ADHD, nicotine use, including vaping, was significantly more prevalent among individuals with a prior ADHD diagnosis, though not with current symptom severity. This discrepancy suggests that nicotine use may reflect long-term behavioral coping or self-medication rather than acute symptom expression.

Opposite to expectation, Stigma did not influence a formal ADHD diagnosis in the sample. However, It still imposes a critical barrier to care as individuals with active ADHD symptoms were significantly more likely to report social stigma as an obstacle to help-seeking. In Jordan and the wider Arab region, where mental health stigma remains prevalent, this finding carries particular relevance and may partially explain undertreatment.

## 5. Conclusion

In Conclusion, this study identified high rates of probable ADHD among young adults attending universities in Jordan. These findings may reflect limitations inherent to the screening process and highlight the need for improved professional training, particularly within primary care settings. Furthermore, addressing the stigma surrounding ADHD treatment, as well as mental health disorders more broadly, is essential. Such stigma -known to be especially prominent in Arab cultures may exacerbate symptom burden and significantly deter help-seeking behaviours among affected individuals.

Collectively, these findings emphasize the importance of accessible, integrated treatment models that address both core ADHD symptoms and associated psychosocial impairments. Expanding access to psychotherapy, academic accommodations, and structured social support may substantially improve functional outcomes for individuals with ADHD.

## Data Availability

All relevant data are within the manuscript and its Supporting Information files

